## Supplementary material for "Prediction of radiation-induced hypothyroidism using radiomic data analysis does not show superiority over standard normal tissue complication models": Fig_S3_Features_radiomic-clinical_models.pdf

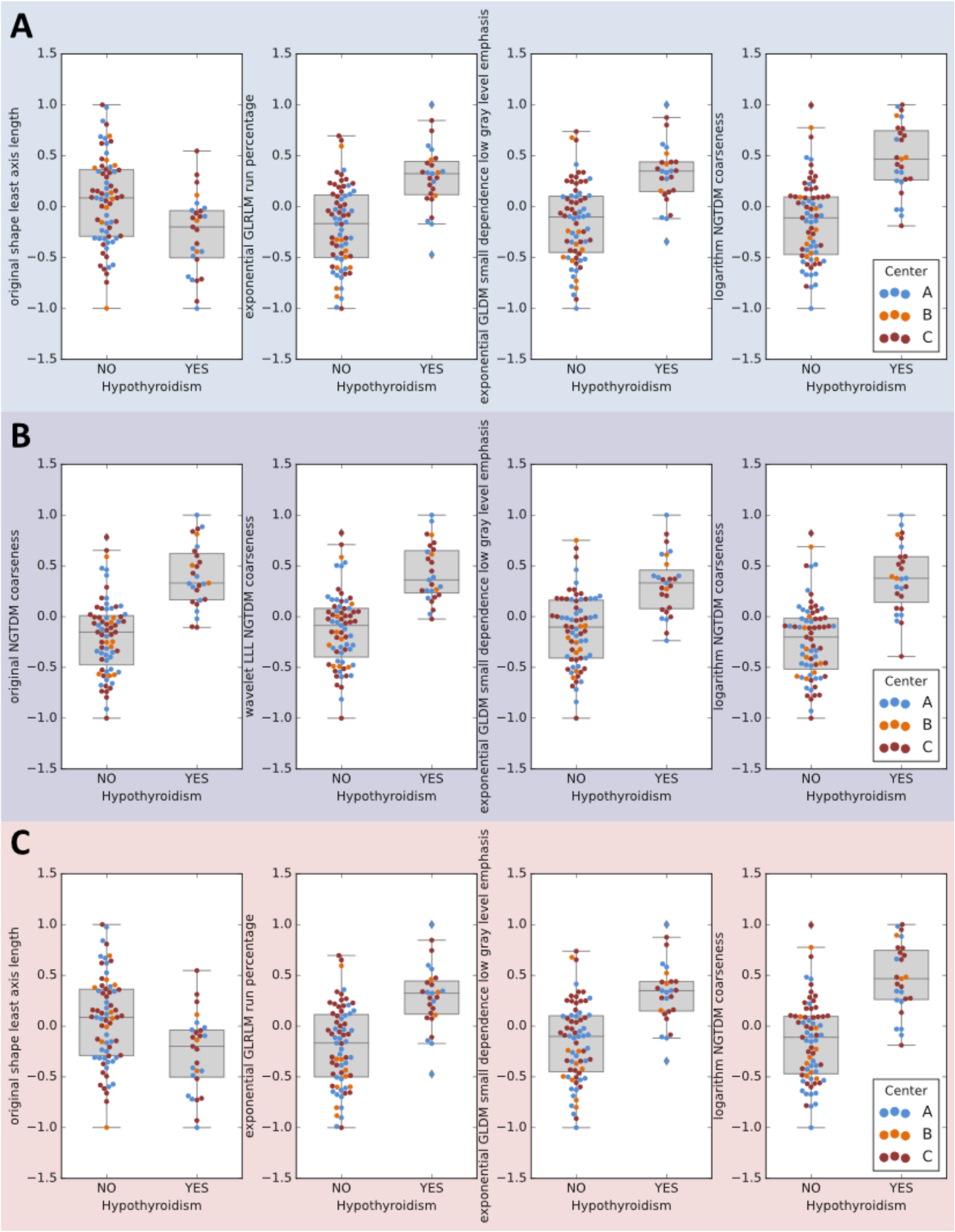


Suppl. Fig. 3: Transformed values of radiomic features retained in radiomic-clinical models. A: variant Ia, B: variant: Ib, C: variant II.
