## Supplementary material for "Prediction of radiation-induced hypothyroidism using radiomic data analysis does not show superiority over standard normal tissue complication models": Table_S3_Models_parameters.pdf

Suppl. Table 2: Tested model architectures and parameters

| LOGISTIC REGRESSION | | | |
|---|---|---|---|
| **Symbol** | **Regularization** | **Class weights** | **Intercept** |
| $LR_B$ | no regularization | balanced | intercept excluded |
| $LR_{BI}$ | | | intercept included |
| $LR_E$ | | equal | intercept excluded |
| $LR_{EI}$ | | | intercept included |

| MULTILAYER PERCEPTRON (MLP) | | |
|---|---|---|
| **Symbol** | **Training parameters** | **Hidden layer(s) size(s)** |
| $MLP_2$ | learning rate: 0.1<br>no regularization<br>activation function: logistic<br>max. iteration number: 1000 | 2 |
| $MLP_3$ | | 3 |
| $MLP_4$ | | 4 |
| $MLP_5$ | | 5 |
| $MLP_{3,3}$ | | 3, 3 |
| $MLP_{4,2}$ | | 4, 2 |

| K NEAREST NEIGHBORS CLASSIFIER | |
|---|---|
| **Symbol** | **Number of neighbors** |
| $KNN_3$ | 3 |
| $KNN_5$ | 5 |
| $KNN_7$ | 7 |

| SUPPORT VECTOR CLASSIFIER (SVC) | | | |
|---|---|---|---|
| **Symbol** | **Class weights** | **Kernel** | **Probability estimation** |
| $SVC_B$ | balanced | RBF | enabled |
| $SVC_E$ | equal | RBF | enabled |

| DECISION TREE | |
|---|---|
| **Symbol** | **Class weights** |
| $DT_B$ | balanced |
| $DT_E$ | equal |

| RANDOM FOREST | | |
|---|---|---|
| **Symbol** | **Class weights** | **Number of decision trees** |
| $RF_{3B}$ | balanced | 3 |
| $RF_{3E}$ | equal | 3 |
| $RF_{5B}$ | balanced | 5 |
| $RF_{5E}$ | equal | 5 |

| ADA BOOST CLASSIFIER | | |
|---|---|---|
| **Symbol** | **Basic estimator type** | **Number of estimators** |
| $AB_3$ | decision tree | 3 |
| $AB_5$ | decision tree | 5 |

| OTHER MODELS | | |
|---|---|---|
| **Symbol** | **Model type** | **Parameters** |
| GPC | Gaussian process classifier | |
| GNB | Gaussian Naive Bayes classifier | priors=[0.5, 0.5] |
| QDA | Quadratic discriminant analysis | priors=[0.5, 0.5] |
