## Supplementary figures and images for "Prediction of radiation-induced hypothyroidism using radiomic data analysis does not show superiority over standard normal tissue complication models"

### Fig_S1_Normalization_scaling.pdf

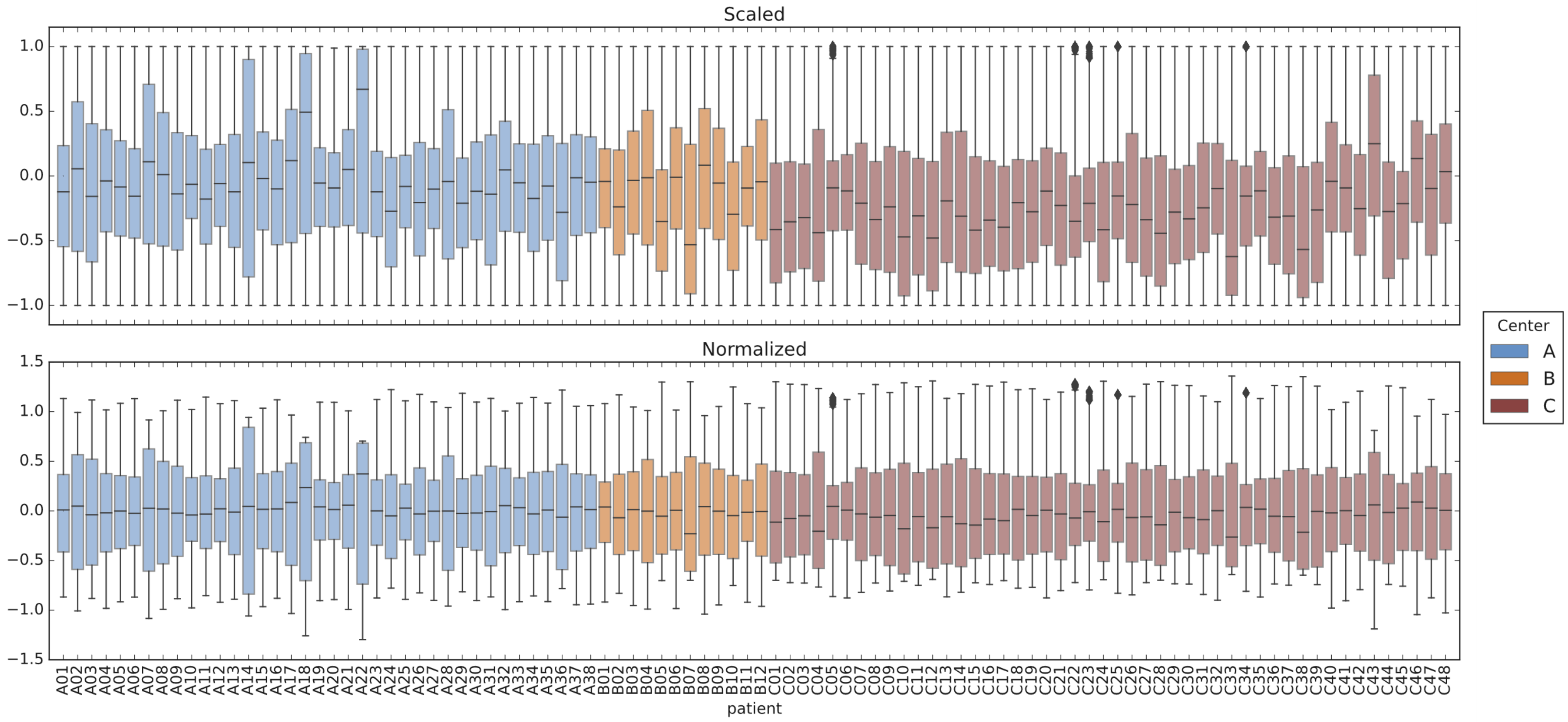


Fig. S1: Scaling and normalization of radiomic features' values.

### Fig_S2_Batch_effect_correction.pdf

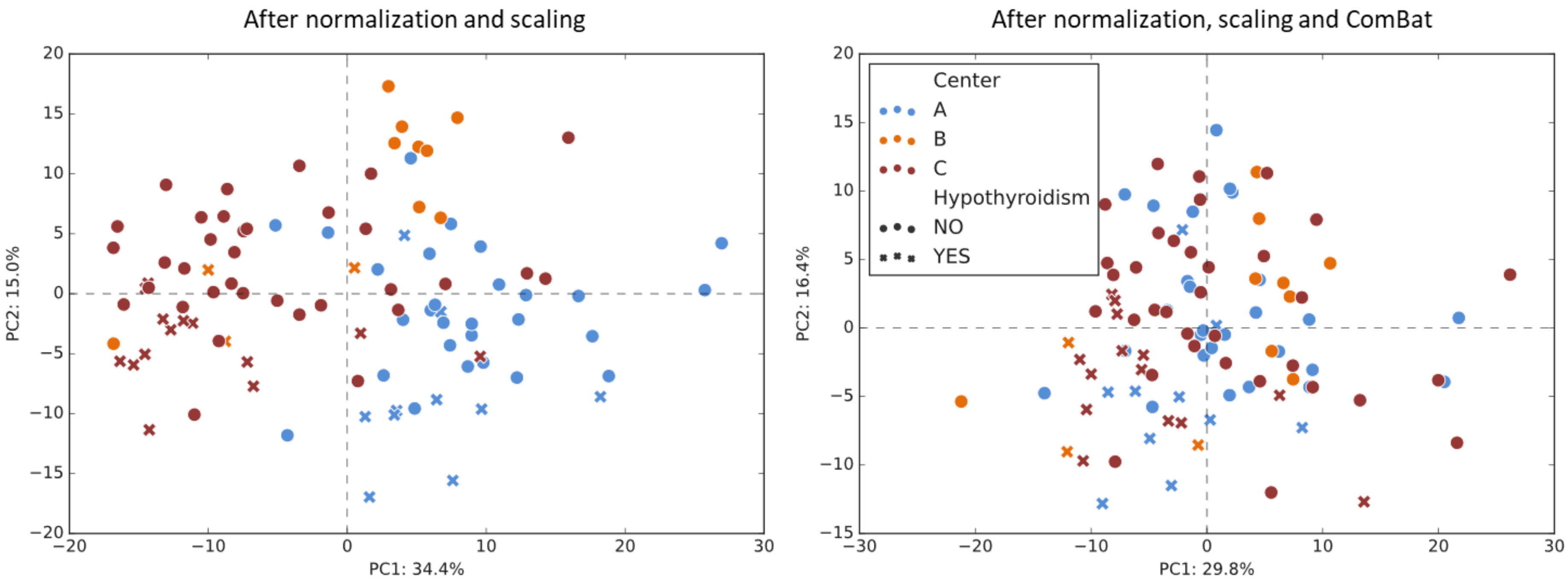


Fig. S2: Principial component analysis before and after batch effect correction.
